# Randomized, Comparative, Clinical Trial to Evaluate Efficacy and Safety of PNB001 in Moderate COVID-19 Patients

**DOI:** 10.1101/2021.04.16.21255256

**Authors:** Eric Lattmann, Pradnya Bhalerao, BL ShashiBhushan, Neeta Nargundkar, Pornthip Lattmann, K Sadasivan Pillai, PN Balaram

## Abstract

**Objective:** To evaluate the efficacy and safety of PNB001 a CCK-A agonist and CCK-B antagonist, a new chemical entity with anti-inflammatory and immune stimulation properties, along with Standard of Care (SOC) in patients with moderate COVID-19 infection.

**Design:** Multi-center, randomized, parallel group, comparative, open label study.

**Setting:** Two tertiary-care hospitals in India.

**Participants:** Patients with laboratory-confirmed SARS-CoV-2 infection as determined by polymerase chain reaction (PCR) within 2 days of randomization, having pneumonia with no signs of severe disease (severe disease means SpO2≤94% on room air), and any two of the following signs or symptoms suggestive of COVID-19: fever, cough, dyspnea, or hypoxia.

**Interventions:** Patients were randomized 1:1 to receive PNB001 at an oral dose of 100 mg three times daily for 14 days with Standard of Care (PNB001+SOC) or only SOC.

**Main outcome measures:** The primary endpoints were mean change in the 8-point WHO Ordinal Scale score from baseline by Day 14 and mortality rate by Day 28. The key secondary endpoints were percentage of patients showing change in clinical status using the ordinal scale, improvement in inflammatory segments in X-ray chest, reduction of days of hospitalization, duration of supplemental oxygen use, days to negative PCR for COVID-19 and change in inflammation markers Interlukin-6 (IL6) and C-reactive protein (CRP) from baseline by Day 14.

**Results:** A total of 40 (20 in PNB 001+SOC arm and 20 in SOC arm) patients were randomized and received treatment. The primary endpoint showed significant clinical improvement from baseline to Day 14 with PNB001+SOC (0.22 Vs 1.12; P=0.0421). One patient in PNB001+SOC arm and two patients in SOC arm died (1 Vs 2; HR: 2.0 [95%CI=0.18, 22.05]; P=0.5637) by Day 28. At the end of the treatment by Day 14, more patients achieved zero ordinal scale in PNB001+SOC arm (17 Vs 12; P=0.0766). In the PNB001+SOC arm, change in mean chest X-ray score showed significant improvement (2.05 Vs 1.16; P=0.0321), and more patients quickly showed complete improvement (10 Vs 7; HR: 1.48 [95%CI=0.64, 3.44]; P=0.4309). In the PNB001+SOC arm, patients needed shorter duration of hospitalization in days (9.45 Vs 9.80) and more patients attained earlier discharge from the hospital (19 Vs 15; P=0.0486) with respect to days. The mean duration of supplemental oxygen requirement in days was shorter (5.45 Vs 7.10) and complete withdrawal from supplemental oxygen was more frequent with PNB001+SOC compared to SOC by Day 14 (17 Vs 13; P=0.1441). All patients in both the arms had negative PCR by the end of the study (18 Vs 17; P=0.6265) by similar time (7.6 Vs 7.0). Exploratory analysis done for IL-6, CRP, Neutrophil-Lymphocyte-Ratio (NLR), Platelet-Lymphocyte-Ratio (PLR) and Erythrocyte Sedimentation Rate (ESR) showed statistically significant reduction by Day 14 demonstrating PNB001’s anti-inflammatory and immunomodulatory properties. Lymphocyte and neutrophil counts also improved by Day 14. 11 adverse events (AE) in 8 patients were observed with PNB001+SOC compared to 13 AEs in 10 patients with SOC; none of the AEs in PNB001+SOC arm were related to PNB001. The most common AE were tachycardia and acute respiratory distress syndrome; there were isolated cases of hepatic enzyme elevation and hyperglycemia. Overall, safety profile was similar between PNB001+SOC and SOC arms.

**Conclusions:** PNB001 with standard of care showed significant clinical improvement in moderate COVID-19 patients when compared to standard of care and was well tolerated by moderate COVID-19 patients.

**Trial Registration:** CTRI/2020/10/028423

## INTRODUCTION

Coronavirus disease 2019 (COVID-19) has widely spread over the globe since its first case in December 2019 in Wuhan, China. COVID-19 is declared as a pandemic by the WHO.^1^ COVID-19 is spread via droplets or direct contact and infects the respiratory tract resulting in pneumonia in most of the cases and acute respiratory distress syndrome (ARDS) in about 15% of the cases.^2^

During COVID-19 infection, a large amount of pro-inflammatory cytokines gets released resulting in an aggressive inflammatory response. The event of gust of pro-inflammatory cytokines is known as “cytokine storm”, and it shows a direct relation with the increased mortality in COVID-19.^2^ High levels of IL-6, C-reactive protein (CRP), neutrophil-Lymphocyte ratio (NLR), platelet-lymphocyte ratio (PLR), erythrocyte-lymphocyte ratio (ESR), and ferritin are associated with more severe disease.^3–10^ Excessive pro-inflammatory cytokines aggravates ARDS and cause tissue damage leading to multiple organ failure, and unfavorably affect prognosis of severe COVID-19. Severe COVID-19 patients develop cytokine storm syndrome, acute respiratory distress syndrome, organ dysfunction, and death. Neutrophil extracellular traps (NETs) are described as important mediators of tissue damage in inflammatory diseases, and is relevant in COVID-19 pathophysiology.^11^ NLR is a predictive biomarker of serious COVID-19 infection. At advanced stage of COVID-19, overproduction of neutrophils via NETosis leads to “cytokine storm”, and it is directly linked with high mortality.^2^ COVID-19 is an out of tune of the immune system and the main treatment objective is to bring neutrophils and lymphocytes levels back into the healthy range. A therapeutic agent that targets cytokines could improve survival rates and reduce mortality.^2,10^

PNB001, a new chemical entity, is a first in class cholecystokinin-A (CCK-A) agonist and CCK-B antagonist, a unique anti-inflammatory agent with immune stimulation properties. It has shown analgesic, antipyretic and anti-inflammatory properties in animal models.^12,13^ The inflammatory immune response of PNB001 was studied in Dengue infected mice and IL-6 levels were measured. After infection, the levels of pro-inflammatory cytokines were increased until day 3 and levels returned back to normal on week 2 when the virus infection was overcome by the host immune system. PNB001 resulted in a reduction of inflammatory marker IL-6 by 80%, in line with smaller spleen inflammation and a reduced death rate.^14^

PNB001 has the potential to reduce the pro-inflammatory cytokines, and in turn the cytokine storm which is the main cause of fatalities in COVID-19 patients. It acts on the inflammatory cytokines by the cholinergic anti-inflammatory pathway and the gastrin-releasing peptide receptor pathway. It is a dual action CCK receptor ligand acting on the CCK-B / gastrin as antagonist and CCK-A agonist. The cholecystokinin via the CCK-2 pathway, reduces inflammation in addition to the CCK-1 cholinergic anti-inflammatory pathway. Figure 1 depicts the role of cholecystokinin in the immune system.

**Figure 1.**
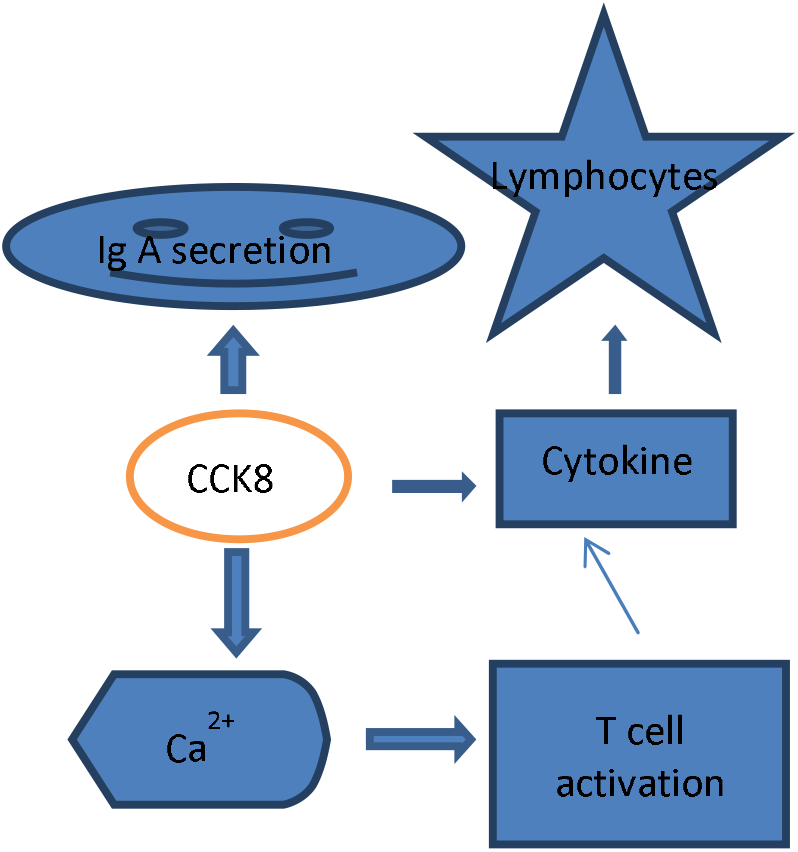
MOA: CCK mediated immunology. Immune regulation: High expression of the cholecystokinin receptor (CCK-A and CCK-B) on B lymphocytes in mouse at mRNA and protein levels. In the spleen CCK receptor is found in high abundance, biologically CCK-8 is supposed a stimulating factor in Ig A and Ig G and IgM to a lesser degree. Key is the increase of the calcium concentration, measured by fura-2 fluorometry, in form of second messengers by the G_q_ pathway (G protein coupled receptor, GPCR).

In a single ascending dose study, PNB001 was administered under fasting condition. The PK (C_max_ and AUC_0-t_) of PNB001 over the dose range of 25-1500 mg was assessed.^15^ In a multiple ascending dose study (50, 100 and 200 mg daily for 14 days), mean half-lives of PNB001 ranged from 2.4 to 6.8 hours on Day 1 and from 5.3 to 7.9 hours on Day 14.^16^ PNB001 was well tolerated in both the studies without any incidence of death or serious adverse events. A 100 mg dose was chosen for the next phase of the study.^15,16^

The objectives of this clinical trial were to assess efficacy and safety of PNB001 in combination with standard of care (SOC) in patients with moderate COVID-19 Infection and to compare it with SOC alone.

## METHODS

### Study Design and Conduct

This was a multi-center, randomized, parallel group, comparative, open label, clinical study to evaluate efficacy and safety of PNB001 in patients with moderate COVID-19 infection.

The study was conducted in compliance to International Council for Harmonisation (ICH) guidance for Good Clinical Practice (GCP) and CDSCO (Schedule-Y) guidance of New Drugs and Clinical Trials Rules, 2019 (India). The study protocol was approved by independent ethics committee. All the patients provided written informed consent prior to study enrolment. The study was conducted at two sites in India (Study Registration No. CTRI/2020/10/028423).

### Study Population

The inclusion criteria of the study were laboratory-confirmed SARS-CoV-2 infection as determined by PCR within 2 days of randomization; patients having pneumonia with no signs of severe disease (severe disease means SpO2 ≤94%) on room air; patients with any two of the following signs or symptoms suggestive of COVID-19: fever, cough, dyspnea, or hypoxia; respiratory rate more or equal to 24 per minute; radiographic infiltrates as confirmed by imaging (chest X-ray). Patients were excluded if they required invasive mechanical ventilation; had the following clinically significant laboratory abnormalities: SGOT, SGPT, serum bilirubin > 2.5 times the Upper Limit Normal (ULN); had abnormal serum creatinine value of ≥ 2 mg/dl; had Type 1 diabetes mellitus or uncontrolled Type 2 diabetes mellitus with random sugar ≥ 200 mg/dL; or had uncontrolled hypertension (systolic blood pressure> 160 mm Hg, or diastolic blood pressure>100mmHg) or previous history of hypertension crisis or hypertensive encephalopathy.

### Randomization and Interventions

Patients were randomized in 1:1 ratio to receive either PNB001 along with Standard of Care (PNB001+SOC) or only Standard of Care (SOC). Patients, who were randomized to PNB001+SOC arm, received 100 mg capsule of PNB001 orally thrice daily for 14 days along with SOC. Patients randomized to other arm received only SOC. Standard of Care was considered as per Clinical Management Protocol: COVID-19 Ministry of Health and Family Welfare Directorate General of Health Services, India and PI discretion.

The site team recorded the details of the study medication administration in the drug dosing card including date, time, dose, frequency, and missed dose information. During the treatment period of 14 days, patients were assessed daily for the improvement in clinical signs and symptoms, concomitant medications, and AE. Patients were withdrawn if they withdrew consent, developed adverse event requiring discontinuation of medication or needed prohibited medications, was not responding or condition was worsening on treatment or SpO2 fell below 90% on room air during the trial or if investigators concluded that for safety or tolerability reasons it is best for the patient to be withdrawn from the trial. Withdrawn patients received other effective therapy, if available.

### Study Endpoints

Primary endpoints of the study were change in the 8-point WHO Ordinal Scale score for COVID-19 from baseline to Day 14 and mortality rate by Day 28. Secondary endpoints were (i) percentage of patients showing change in clinical status using the ordinal scale from baseline; (ii) percentage of patients showing improvement in inflammatory segments in X-ray chest from baseline; (iii) reduction of days of hospitalization; (iv) duration of supplemental oxygen (if applicable) (v) improvement in oxygen saturation from baseline; and (vi) days to negative PCR for COVID-19 and (vii) change in inflammation markers IL6 and CRP from baseline to Day 14. The endpoint -improvement in oxygen saturation from baseline was later removed because when condition of the patient worsened, another mode of oxygen administration was followed, making oxygen saturation comparison, not a useful parameter.

Exploratory analysis was done for IL-6, CRP, NLR, PLR, and ESR to assess mean change in these markers from baseline to Day 14.

### Safety Assessments

The assessment of safety of PNB001 was based on the frequency of AEs, changes in laboratory values and abnormal vital signs. Safety laboratory parameters were complete blood counts, erythrocytes sedimentation rate, liver function tests, total bilirubin, SGOT, SGPT, serum creatinine, CRP, and IL6 with X-ray Chest and ECG. Vital signs were measured at all visits.

### Statistical Analysis

Continuous efficacy and safety variables were presented using descriptive statistics which included number, mean, standard deviation, median, and range. Categorical data were presented using counts and percentages. Mean change in the ordinal scale from baseline to end of study was analyzed using paired Wilcoxon signed rank test and 2 sample Wilcoxon rank sum tests. Mortality through Day 28 were analyzed as a time to event endpoint. Differences in time-to-event endpoints by treatment were summarized with Kaplan-Meier curves. Secondary efficacy parameters were analyzed and presented using descriptive statistics. Changes in the inflammatory markers (IL-6, CRP) from baseline to end of study were analyzed using paired Wilcoxon signed rank test and 2 sample Wilcoxon signed rank tests. Exploratory comparative analysis for IL-6, CRP, NLR, PLR and ESR were done within group using paired Wilcoxon rank sum test and between groups using Wilcoxon rank sum test.

All the statistical analyses were performed using SAS 9.2 or higher version software.

### Patient and public involvement

Patients were not involved in the decision of treatment choice, study design, conduct, and interpretation or reporting of results of this study.

## RESULTS

### Study Participants

A total of 42 patients were screened of which one failed during screening and one withdrew consent before randomization. Hence, 40 patients were randomized into two arms, i.e. 20 patients/treatment arm. In PNB001+SOC arm, 18/20 patients completed the study; one patient withdrew consent and one patient was discontinued from the study at the discretion of investigator, as the disease worsened in this patient. In SOC arm, 17/20 patients completed the study; one patient withdrew consent and two patients discontinued from the study at the discretion of investigator as the disease worsened. Thus, total of 35 patients completed the study. The patients whose disease worsened and were discontinued by the investigator, subsequently, succumbed to the disease. Figure 2 depicts the patient disposition flow.

**Figure 2.**
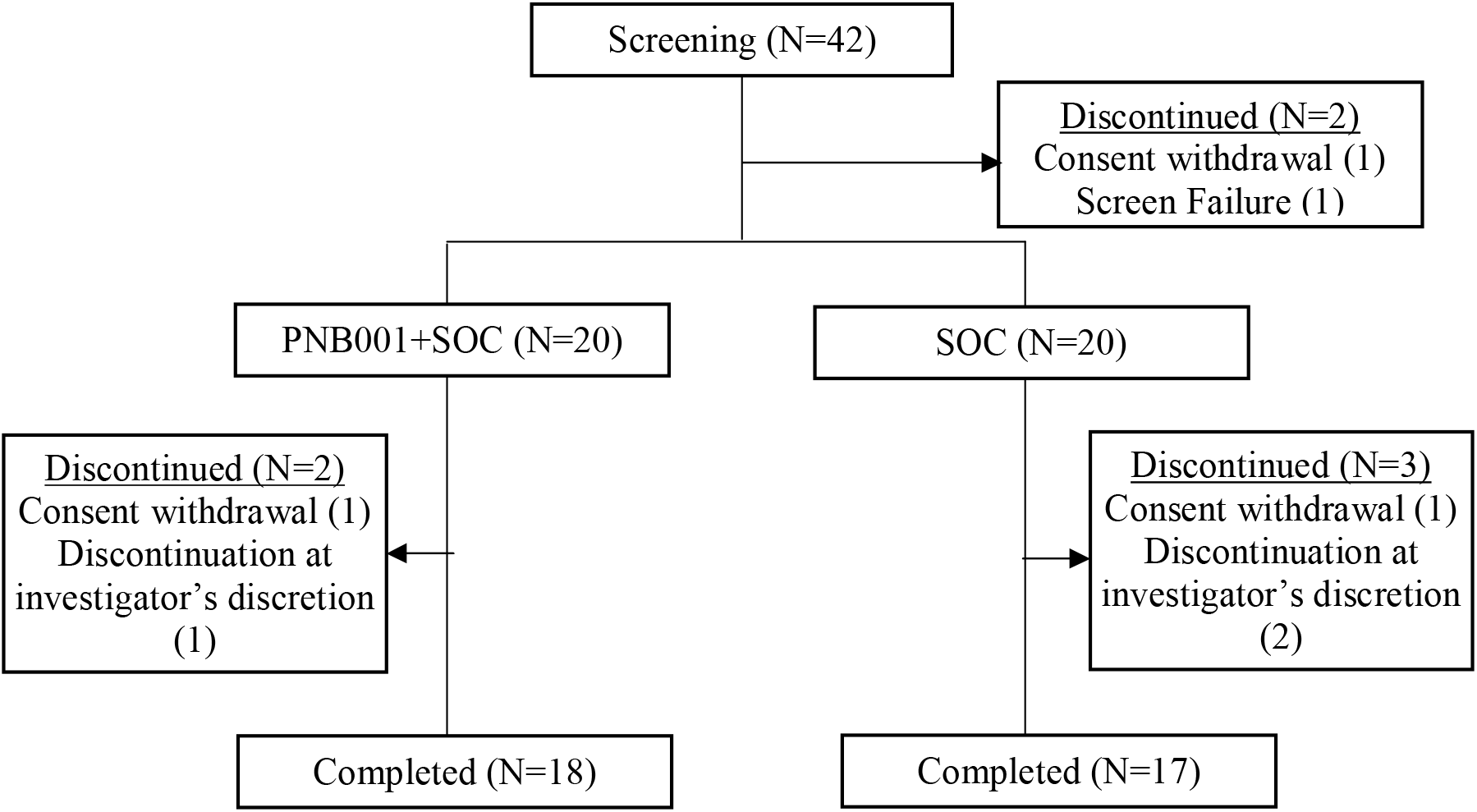
Patient Disposition by Treatment Arms. SOC=Standard of care

### Patients Demography and Baseline Characteristics

There were 14 males in the PNB001+SOC arm and 12 males in the SOC arm. The mean patient age (52.10 years) in PNB001+SOC arm was comparable with (52.8 years) SOC arm. The mean body height (158.30 cm) in PNB001+SOC arm was comparable with (156.0 cm) SOC arm. The average body weight (61.90 kg) in PNB001+SOC arm was comparable with (56.75 kg) SOC arm. All patients were Indian. The demographics of patients were balanced between two treatment arms (Supplementary Table 1).

Two patients in PNB001+SOC arm had history of tobacco use; one was an occasional user and other consumed 10-15 packs of tobacco every week. The concomitant medication usage was comparable between the treatment arms. Commonly used concomitant medication were remdesivir, steroids, heparins, aspirin, different antibiotics, ivermectin, zinc, vitamin C, drugs for acidity, antiemetics, etc. A few patients had comorbid disease such as controlled diabetes mellitus (7), asthma (1), hypertension (6), hypothyroidism (1), deep vein thrombosis (2), pulmonary embolism (1), hyperglycaemia (1) and obesity (1). The baseline characteristics of patients were comparable between the treatment arms (Supplementary Table 1).

### Efficacy Outcomes

#### Primary efficacy endpoints

Patients in PNB001+SOC (test) arm showed significant improvement in WHO Ordinal Scale for COVID-19 Clinical Improvement compared to patients in SOC (control) arm at the end of the treatment (Figure 3). Further, patients showed significant Clinical improvement from baseline from Day 5 onwards till end of the study in PNB001+SOC arm; patients who received only SOC showed significant improvement from baseline from Day 8 onwards. At the end of the treatment (Day 14), mean ordinal scale was 0.22 in PNB001+SOC arm and 1.12 in SOC arm, and the mean change in ordinal scale from baseline to Day 14 was statistically significant (P=0.0421).

**Figure 3.**
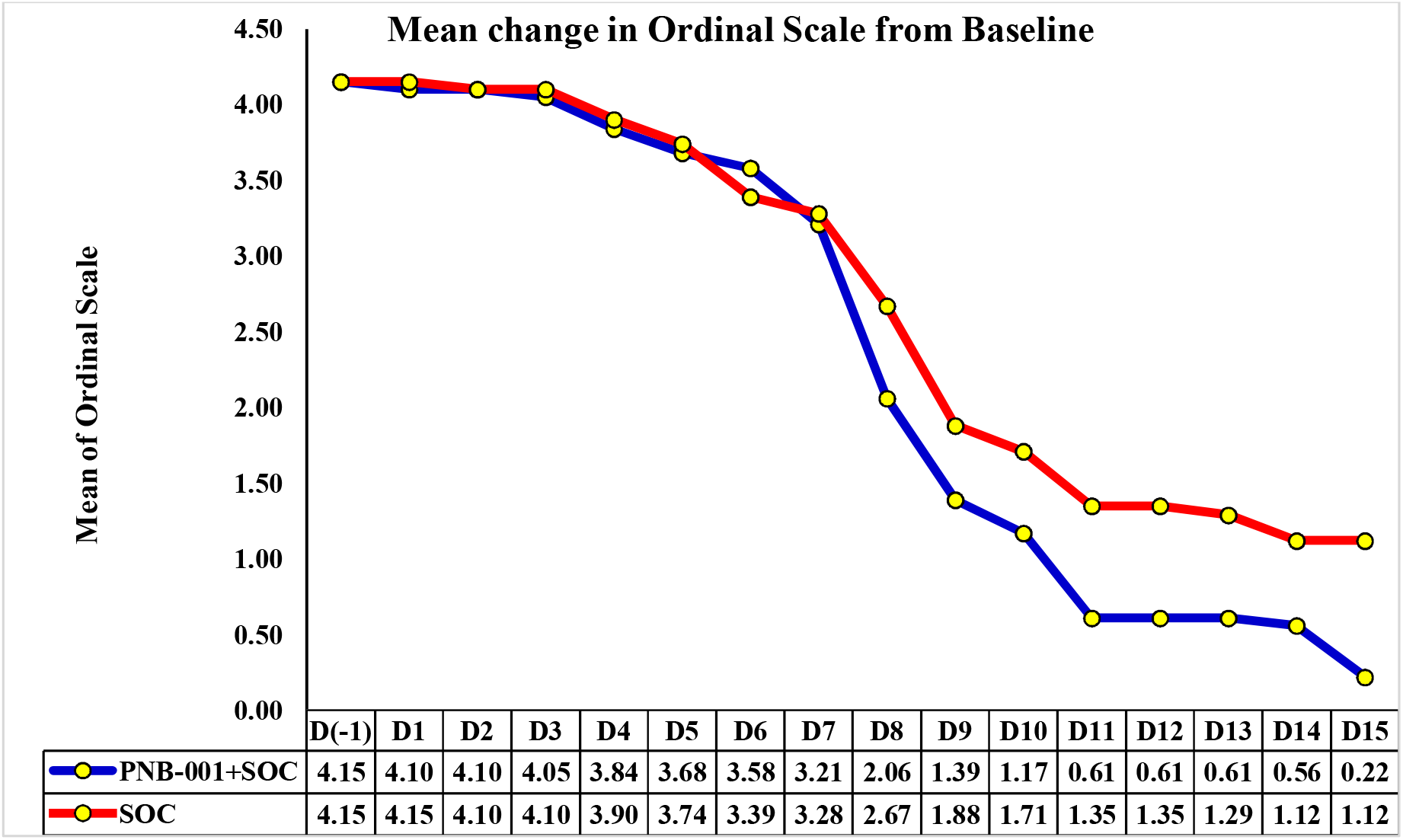
Ordinal Scale Mean change from Baseline. SOC=Standard of Care

During the study, one patient in PNB001+SOC arm died on Day 4 after receiving PNB001 for only 2 and half days. Two patients died in SOC arm; one patient on Day 6, the other patient on Day 10. These patients were discontinued from the study by the investigator as per the protocol due to worsening of clinical condition, prior to the event. Mortality rate was comparable between the test and control arms (HR: 2.0 [95%CI=0.18, 22.05]; P=0.5637), though numerically it favored test arm.

#### Secondary efficacy endpoints

A summary of change in ordinal scale from baseline to all the visits is presented in Table 1. Higher number of patients achieved ordinal scale of zero in PNB001+SOC arm compared to SOC arm from Day 8 onwards. At the end of the treatment, 17 (94.44%) patients achieved zero ordinal scale in PNB001+SOC arm, whereas, 12 (70.59%) patients achieved zero ordinal scale in SOC arm (P=0.0766). Other secondary efficacy results are presented in Table 2. X-ray assessment of inflammatory segments of lungs showed improvement in higher number of patients in PNB001+SOC arm compared SOC arm (50.00% vs. 36.84%, HR:1.48 [95% CI: 0.64,3.44]; P=0.4309). Mean change in the chest X-ray score was 2.05 (SD=1.19) in PNB001+SOC arm and 1.16 (SD=1.21) in SOC arm (P=0.0321). Patients receiving PNB001+SOC were hospitalized for lesser duration compared to SOC (Mean 9.45 vs. 9.80 days). Significantly higher number of patients were discharged from the hospital by Day 14 in PNB001+SOC arm compared to SOC arm (19 vs. 15, P=0.0486). Mean duration of requirement of supplemental oxygen was shorter for patients in the test arm (5.45 vs. 7.10 days). In PNB001+SOCarm, 17 (94.44%) patients were off Oxygen support on Day 14 whereas only 13 (76.47%) patients were off Oxygen support with SOC (P=0.1441). A summary of different methods of oxygen administered during the study is provided in Supplementary Table 3. PCR test was negative by median of Day 8 for both the treatment arms (P=0.6265).

**Table 1:** Summary of Ordinal Scale from Baseline to End of Day 14.

| Visit | Scale | PNB001 (N=20)<br>n (%) | Standard of Care (N=20)<br>n (%) |
| --- | --- | --- | --- |
| **Baseline** | 4-Oxygen by mask or nasal prongs | 17 (85.00%) | 17 (85.00%) |
|  | 5-Non-invasive ventilation or high flow oxygen | 03 (15.00%) | 03 (15.00%) |
| **Day 1** | 4-Oxygen by mask or nasal prongs | 18 (90.00%) | 17 (85.00%) |
|  | 5-Non-invasive ventilation or high flow oxygen | 02 (10.00%) | 03 (15.00%) |
| **Day 2** | 3-Hospitalized, no oxygen therapy | 00 (00.00%) | 02 (10.00%) |
|  | 4-Oxygen by mask or nasal prongs | 18 (90.00%) | 14 (70.00%) |
|  | 5-Non-invasive ventilation or high flow oxygen | 02 (10.00%) | 04 (20.00%) |
| **Day 3** | 3-Hospitalized, no oxygen therapy | 02 (10.00%) | 03 (15.00%) |
|  | 4-Oxygen by mask or nasal prongs | 15 (75.00%) | 12 (60.00%) |
|  | 5-Non-invasive ventilation or high flow oxygen | 03 (15.00%) | 05 (25.00%) |
| **Day 4** | 0-No clinical or virological evidence of infection | 00 (00.00%) | 01 (05.00%) |
|  | 3-Hospitalized, no oxygen therapy | 05 (26.32%) | 03 (15.00%) |
|  | 4-Oxygen by mask or nasal prongs | 12 (63.16%) | 11 (55.00%) |
|  | 5-Non-invasive ventilation or high flow oxygen | 02 (10.53%) | 05 (25.00%) |
| **Day 5** | 0-No clinical or virological evidence of infection | 00 (00.00%) | 02 (10.53%) |
|  | 3-Hospitalized, no oxygen therapy | 08 (42.11%) | 02 (10.53%) |
|  | 4-Oxygen by mask or nasal prongs | 09 (47.37%) | 10 (52.63%) |
|  | 5-Non-invasive ventilation or high flow oxygen | 02 (10.53%) | 05 (26.32%) |
| **Day 6** | 0-No clinical or virological evidence of infection | 00 (00.00%) | 03 (16.67%) |
|  | 3-Hospitalized, no oxygen therapy | 10 (52.63%) | 02 (11.11%) |
|  | 4-Oxygen by mask or nasal prongs | 07 (36.84%) | 10 (55.56%) |
|  | 5-Non-invasive ventilation or high flow oxygen | 02 (10.53%) | 03 (16.67%) |
| **Day 7** | 0-No clinical or virological evidence of infection | 01 (05.26%) | 03 (16.67%) |
|  | 3-Hospitalized, no oxygen therapy | 13 (68.42%) | 04 (22.22%) |
|  | 4-Oxygen by mask or nasal prongs | 03 (15.79%) | 08 (44.44%) |
|  | 5-Non-invasive ventilation or high flow oxygen | 02 (10.53%) | 03 (16.67%) |
| **Day 8** | 0-No clinical or virological evidence of infection | 07 (38.89%) | 05 (27.78%) |
|  | 3-Hospitalized, no oxygen therapy | 08 (44.44%) | 05 (27.78%) |
|  | 4-Oxygen by mask or nasal prongs | 02 (11.11%) | 07 (38.89%) |
|  | 5-Non-invasive ventilation or high flow oxygen | 01 (05.56%) | 01 (05.56%) |
| **Day 9** | 0-No clinical or virological evidence of infection | 11 (61.11%) | 08 (47.06%) |
|  | 3-Hospitalized, no oxygen therapy | 04 (22.22%) | 04 (23.53%) |
|  | 4-Oxygen by mask or nasal prongs | 02 (11.11%) | 05 (29.41%) |
|  | 5-Non-invasive ventilation or high flow oxygen | 01 (05.56%) | 00 (00.00%) |
| **Day 10** | 0-No clinical or virological evidence of infection | 12 (66.67%) | 09 (52.94%) |
|  | 3-Hospitalized, no oxygen therapy | 04 (22.22%) | 03 (17.65%) |
|  | 4-Oxygen by mask or nasal prongs | 01 (05.56%) | 05 (29.41%) |
|  | 5-Non-invasive ventilation or high flow oxygen | 01 (05.56%) | 00 (00.00%) |
| **Day 11** | 0-No clinical or virological evidence of infection | 15 (83.33%) | 11 (64.71%) |
|  | 3-Hospitalized, no oxygen therapy | 02 (11.11%) | 01 (05.88%) |
|  | 4-Oxygen by mask or nasal prongs | 00 (00.00%) | 05 (29.41%) |
|  | 5-Non-invasive ventilation or high flow oxygen | 01 (05.56%) | 00 (00.00%) |
| **Day 12** | 0-No clinical or virological evidence of infection | 15 (83.33%) | 11 (64.71%) |
|  | 3-Hospitalized, no oxygen therapy | 01 (05.56%) | 01 (05.88%) |
|  | 4-Oxygen by mask or nasal prongs | 02 (11.11%) | 05 (29.41%) |
| **Day 13** | 0-No clinical or virological evidence of infection | 15 (83.33%) | 11 (64.71%) |
|  | 3-Hospitalized, no oxygen therapy | 01 (05.56%) | 02 (11.76%) |
|  | 4-Oxygen by mask or nasal prongs | 02 (11.11%) | 04 (23.53%) |
| **Day 14** | 0-No clinical or virological evidence of infection | 15 (83.33%) | 12 (70.59%) |
|  | 3-Hospitalized, no oxygen therapy | 02 (11.11%) | 01 (05.88%) |
|  | 4-Oxygen by mask or nasal prongs | 01 (05.56%) | 04 (23.53%) |
| **Day 15** | 0-No clinical or virological evidence of infection | 17 (94.44%) | 12 (70.59%) |
|  | 3-Hospitalized, no oxygen therapy | 00 (00.00%) | 01 (05.88%) |
|  | 4-Oxygen by mask or nasal prongs | 01 (05.56%) | 04 (23.53%) |

**Table 2:**
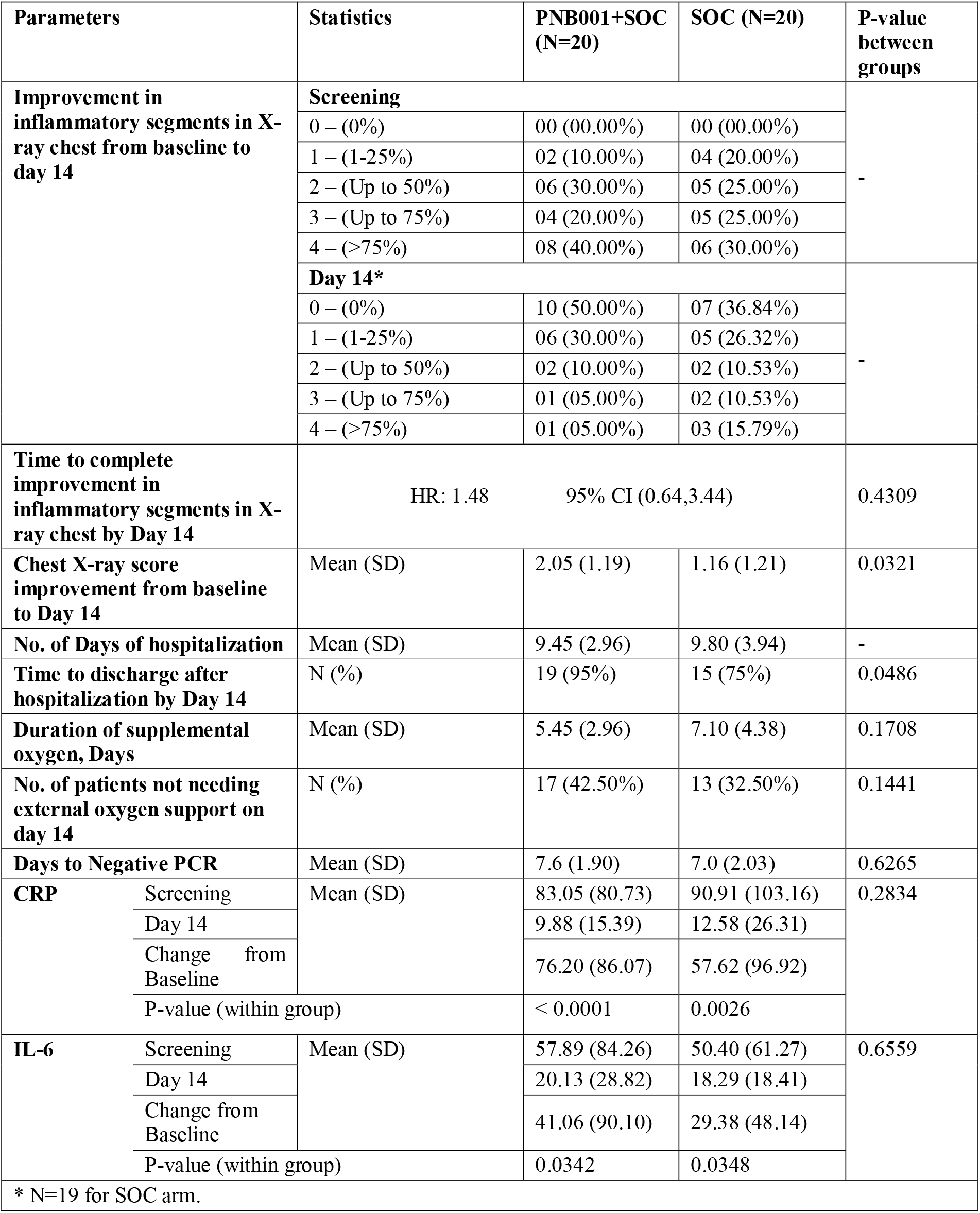
Secondary Efficacy Assessments.

**Table 3:**
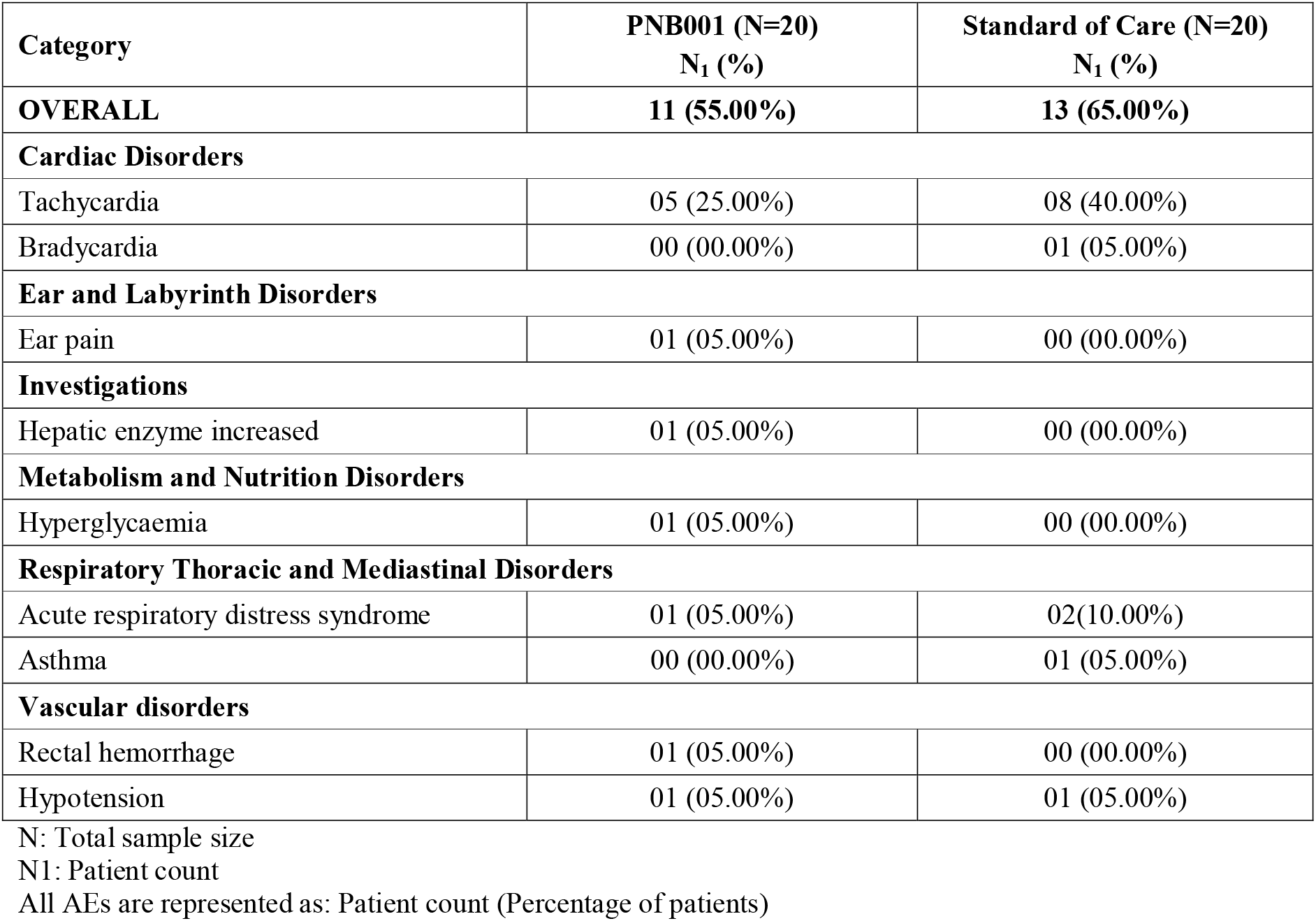
Summary of AEs – Safety Population.

#### Exploratory efficacy endpoints

Exploratory analysis done for IL-6, CRP, NLR, PLR and ESR showed statistically significant reduction in values on Day 14 compared to baseline in both the treatment arms demonstrating PNB001’s anti-inflammatory and immunomodulatory properties. Patients whose values were within the normal range at the beginning of the study were removed and analysis was performed using the remaining patient data (this was to ensure inclusion of patients who had severe infection alone for comparison). The mean decrease of CRP, IL-6, NLR, PLR, and ESR was larger in PNB001+SOC arm compared to SOC arm and these parameters improved significantly by day 14 with both the treatments. However, there was no statistically significant difference between arms (Supplementary Table 4). Figure 4 to 9 depicts effects of PNB001+SOC and SOC on CRP, IL-6, Lymphocyte count, Neutrophil count and glucose level levels.

**Figure 4.**
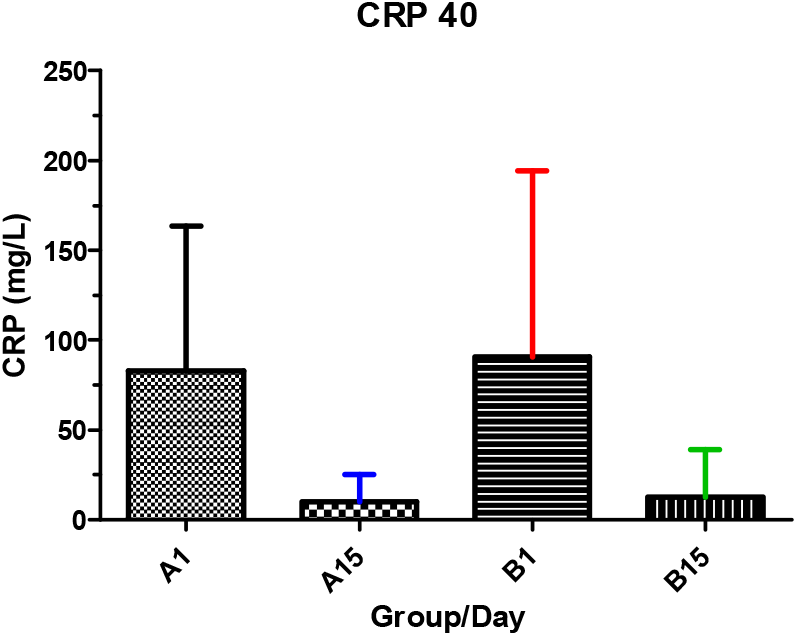
C-reactive Protein A-PNB001+SOC on day 1 and day 15, B-SOC on day 1 and day 15. CRP was reduced in adjunct group by >90% and less than 80% in the best care group. The decrease was larger than the decrease anticipated with any known anti-inflammatory agent. ESR is an inflammation biomarker at the cellular level and CRP an inflammation biomarker at the molecular level. The reference value for CRP for healthy patients is within the range of 1-5 mg/L. At molecular level of 20 mg/ml, inflammation is classified as mild and at 60 mg/ml as moderate respectively.

**Figure 5.**
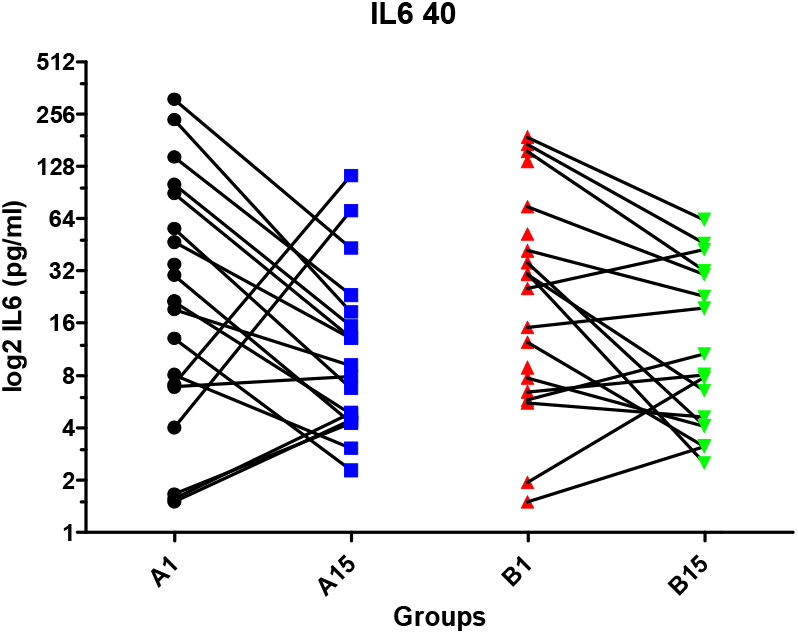
Interleukin-6 A-PNB001+SOC on day 1 and day 15, B-SOC on day 1 and day 15. IL-6 is a key inflammation-/immune bio-marker and is involved in the immune response. Excess cytokines lead to sepsis and organ damage. Organ damage is present in COVID-19 infections. High IL-6 levels in the hundred range are reduced. For high inflammation a reduction of cytokines is observed. For low IL-6 levels in the single digits the increase is observed, leading to the stimulation of the immune system, via CCK-8, starting the proliferation of lymphocytes. IL-6 is an immune marker. Treatment of cancer and viral infection require a full functioning immune system, prior to COVID hypothesis had been postulated with respect to involvement of the immune system.^31^

**Figure 6.**
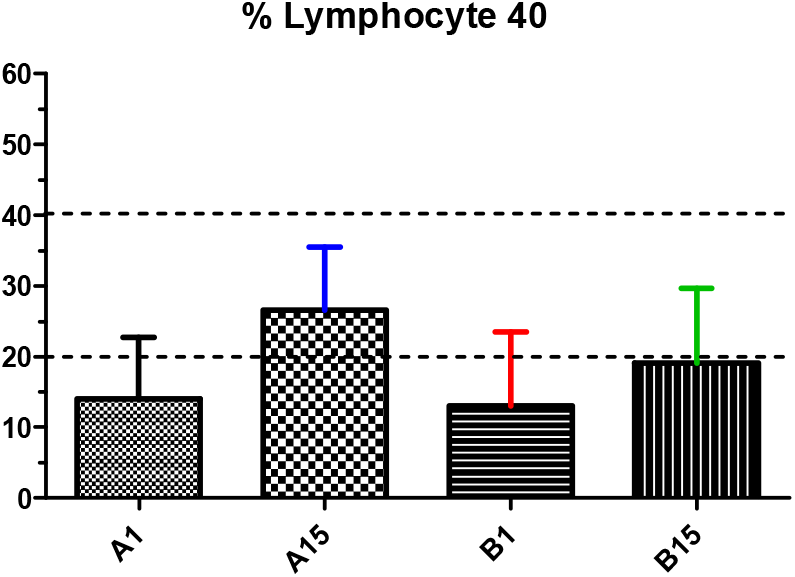
Lymphocyte counts PNB001+SOC on day 1 and day 15, B-SOC on day 1 and day 15. An increase of lymphocytes from 14% to 27% into the middle of the reference range is found for the adjunct treatment after 14 days. For best care the increase is only from 14% to 19%.

**Figure 7.**
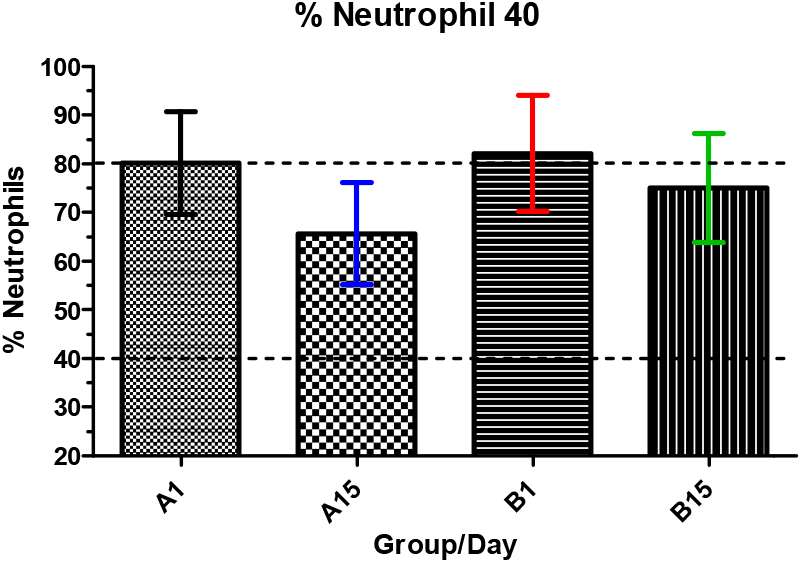
Neutrophil counts A-PNB001+SOC on day 1 and day 15, B-SOC on day 1 and day 15. Reduction of neutrophil is linked with reduction of NEtosis and subsequently prevention of cytokine storm. Here, the number if neutrophils is reduced by the cholinergic inflammatory pathway and the infiltration of neutrophils is reduced by the inhibition of the CCK-B gastrin pathway with involvement of histamine. The percentage of neutrophils was reduced from 81% to 65% on adjunct treatment and from 72% to 76% on best care.

**Figure 8.**
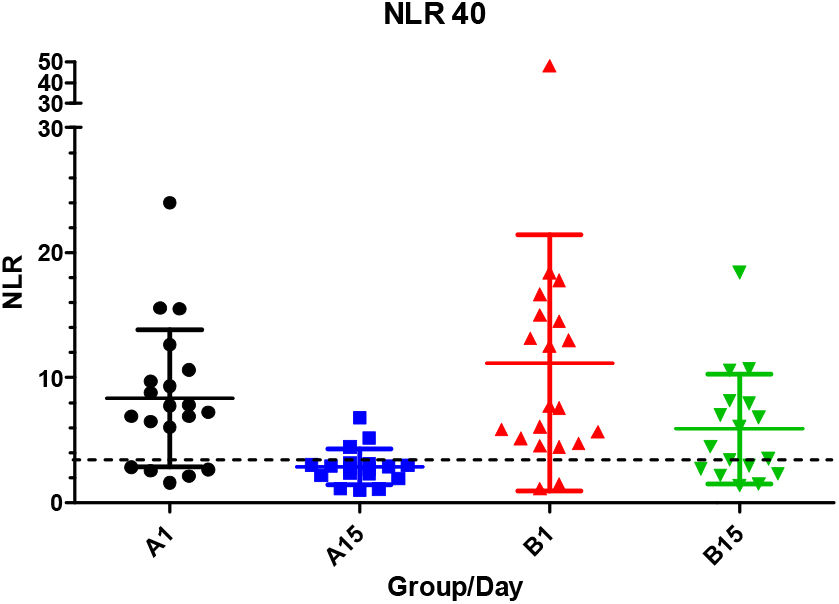
NLR ratio^32^ A-PNB001+SOC on day 1 and day 15, B-SOC on day 1 and day 15. The enrolled patients had an elevated NLR ratio as a result of Covid-19 infection. The NLR was reduced from 8.5 to 2.8 for the adjunct treatment and from 11.1 to 5.9 for best care. The difference between the arms was significant with a P=0.010. A lower than 3.3 ratio was clearly linked with less severe outcome in patients. Here, a reduction of death rate, as well as all other parameters was found. The NLR has been shown to serve as the most reliable indicator of progression to severe COVID-19, as well as a clinical biomarker to monitor treatment. In conclusion, NLR was useful as predictive and as clinical biomarker to assess treatment response.

**Figure 9.**
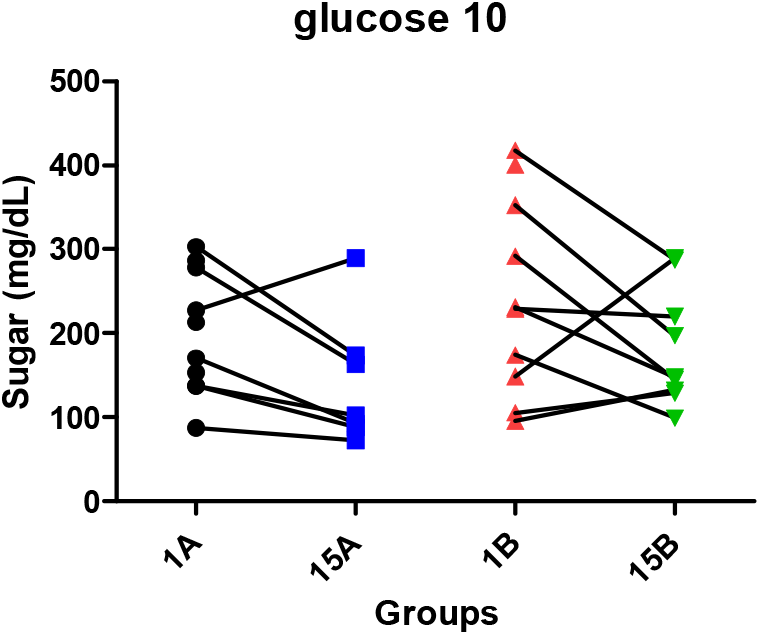
Glucose A-PNB001+SOC on day 1 and day 15, B-SOC on day 1 and day 15. In MAD for the highest dose (200 mg) liver toxicity was observed in one patient (1 in 42). Here, no change of SGOT and SGPT was observed in conjunction with gluco-steroids. For glucose the protocol had a cut-off value of 200 mg/dl. From preclinical experiments, efficacy is higher at higher glucose concentrations and therefore, violation of the protocol was justified with clear clinical benefits to this subgroup. Deviations from the protocol were filed and clearly a reduction of glucose blood concentration was found on adjunct treatment. This is of particular importance as pre-diabetic and diabetic patients have a much worse COVID-19 prognosis and are in much higher treatment need

### Safety Outcomes

A total of 24 AEs were reported by 18 patients during the study: 11 AEs in 8 patients in PNB001+SOC arm and 13 AEs in 10 patients in SOC arm. The adverse events reported were tachycardia, bradycardia, and hypotension, pain in ear, hyperglycaemia, liver enzymes increased, acute respiratory distress syndrome (ARDS), and bleeding per rectum. The most common AEs were tachycardia (5 in PNB001+SOC arm and 8 in SOC arm) and ARDS (1 in PNB001+SOC arm and 2 in SOC arm). Three AEs of ARDS were severe while all other AEs were mild to moderate in severity; none of the AEs were considered related to the study treatments. One SAE of ARDS reported in PNB001+SOC arm was severe though not related to the study treatment; but that resulted in the death of the patient. The death in PNB001 plus standard of care arm was not considered as related to the study treatment.

No clinically significant laboratory abnormality was detected during the study other than an instance of liver enzyme abnormality and another instance of hyperglycaemia documented as adverse event. During vitals assessment, a few events of tachycardia and increased respiratory rate were observed; these events are recorded as AEs. Overall, there was no clinically meaningful difference in the safety profile between PNB001+SOC and SOC arms.

## DISCUSSION

The current COVID-19 pandemic compelled the healthcare industry and other stake-holders to congregate sources to evaluate novel or existing therapeutic intervention for the treatment of COVID-19. The efforts of pharmaceutical companies coalition are swiftly and decisively progressing to provide promising therapeutics agents as an effective treatment option for prevention and treatment of COVID-19. WHO recommends that a randomized controlled trials in combination with supportive care is the suitable way to evaluate the efficacy and safety of treatments for COVID-19.^17^

This randomized study was conducted to assess and compare efficacy and safety of PNB001+SOC with SOC. Patients received 100 mg PNB001 three times a day along with standard of care or only standard of care for 14 days. The study efficacy was assessed based on the primary endpoint of WHO Ordinal Scale for COVID-19 Clinical Improvement besides mortality rate.^18^ The secondary endpoints were the standard endpoints as recommended by WHO and by Natanegara et al.^18,19^

The patient’s demographics, baseline characteristics, and disease severity were comparable between PNB001+SOC and SOC arms.

The primary efficacy endpoint, mean change in ordinal scale, showed significant clinical improvement in PNB001+SOC arm compared to SOC arm on Day 14. Higher number of patients achieved ordinal score zero, i.e. full recovery, in the PNB001+SOC arm from Day 8 to End of the Treatment compared to SOC arm. At the end of the study, all patients in either arms were (94.44% Vs 70.59%) free from clinical or virological evidence of infection by the PCR test. Multiple clinical studies were conducted to assess efficacy of a range of therapeutic agents targeting different mechanism of action for moderate to severe COVID-19 using different ordinal scale methods. Ruiz et al. reported clinical improvement using the 6-pointordinal scale in 44.3% (39/88) and 73.9% (65/88) by days 7 and 14, respectively in severe COVID-19 patients.^20^ Following remdesivir treatment, an ordinal score of 1 was reported in 50.7% (38/75) patients who had baseline ordinal score of 4.^21^

Total three patients (1 in PNB001+SOC arm and 2 in the SOC arm) died during the study; these patients were withdrawn from the study as per the protocol due to worsening of clinical condition prior to the event. The patient who died following randomization to PNB001+SOC is unlikely to have derived benefit from test treatment as half-life of PNB001 is closer to 8 hrs and steady state blood level attainment and pharmacodynamic benefit may require additional days.

The secondary efficacy endpoints, mean Chest X-ray score and time to discharge from the hospital showed statistically significant difference favoring PNB001+SOC arm while other secondary endpoints like number of patients achieving zero ordinal scale, number of patients showing complete Chest X-ray improvement, duration of hospitalization, duration of supplemental oxygen requirement and number of patients who could be totally withdrawn from supplemental oxygen showed more quantifiable improvement in PNB001+SOC arm compared to SOC arm. Days to negative PCR at the end of the study was similar in both the arms. Improvement in oxygen saturation from baseline was planned as an endpoint; however, when oxygen saturation worsened in patients, a more efficient oxygen delivery method was employed making the endpoint less meaningful.

Due to the increased inflammatory processes, NLR and PLR are elevated in COVID-19 patients.^22^ Persistently elevated level of IL-6, ESR, and CRP have been reported in earlier, in severe cases of corona-infected patients.^23^ In the current study, IL6, CRP, NLR, PLR, and ESR were significantly reduced from baseline to Day 14 in both the treatment arm. The reduction in these inflammatory markers was higher in the PNB001+SOC arm compared to SOC arm demonstrating anti-inflammatory and immunomodulatory properties of PNB001; however, the difference between the treatment arms were not significant.

Recent studies on COVID-19 reported NLR as an independent risk factor of the in-hospital death and a significant prognostic biomarker of outcomes in critically ill patients.^24^ In recent years, the bidirectional relationship between the nervous and immune system has become increasingly clear, and its role in both homeostasis and inflammation has been well documented.^25^ Since the introduction of the cholinergic anti-inflammatory pathway, there has been an increased interest in parasympathetic regulation of both innate and adaptive immune responses, including T helper 2 responses.

PNB001 was originally developed for IBD and the link between IBD and immune system is featured.^26^ In vitro, we have shown the release of acetyl choline and acetyl choline regulates immunity.^27^ The subtypes of CCK receptors were discussed and in particular the role of CCK-1, CCK and ACH gives insight in to this. CCK1 receptor activation stimulates vago-vagal reflex pathways in the brain stem.^28^ The cholinergic anti-inflammatory pathway, is the pathway known to modulate cytokine release.

PNB001, is a novel small molecule with CCK-A agonizing and CCK-B antagonizing properties making it a first anti-inflammatory agent analgesic with immune modulating properties. Significantly high blood levels of cytokines and chemokines were noted in patients with COVID-19 infection and these pro-inflammatory cytokines included IL1-β, IL1RA, IL-7, IL-8, IL-9, IL-10, basic FGF2, G-CSF, GM-CSF, IFN-γ, IP10, MCP1, MIP1α, MIP1β, PDGFB, TNF-α, and VEGFA and we consider these cytokines a result of neutrophil overpopulation, NETosis. PNB001 has the potential to reduce the pro-inflammatory cytokines if values are high and to increase cytokines when low measured in form of IL-6 when required to stimulate lymphocyte production. Currently the in-depth analysis of more lymphocytes is performed to understand, if T cells or B cells are formed and which T cells are formed in particular to help fighting the infection. PNB001 acts by stimulation of the immune response indirectly preventing the cytokine storm, which is the main cause of fatality in COVID-19 patients.^29,30^

In the current study, immune inflammatory marker IL-6 was modulated, reduced when too high and increased when low to stimulate to fight of the immune system. The number of lymphocytes is increased and the number of T cells, monitored in form of CD4/CD8 ratio and these results will be reported in due course.

PLR, ESR, killer T-cells, helper T-cells, immunoglobulin IGM and IGG are all involved in an orchestrated response to the COVID-19 infection and are part of PART 2.

PNB001, a first in class CCK-A agonist and CCK-B antagonist with immune stimulation anti-inflammatory properties at a dose of 100 mg thrice daily was well tolerated in patients with moderate COVID-19 infection. Most AEs were mild to moderate in severity and considered not related to the study treatment. There were no abnormal laboratory findings observed during the study other than an instance of liver enzyme abnormality and an instance of hyperglycaemia documented as AEs. Abnormal vitals included tachycardia and increased respiratory rate which were considered as due to disease physiology. No clinically meaningful differences were observed in the safety profiles between PNB001+SOC and SOC treatment arms.

Often COVID 19 infected patients experience lingering symptoms like fatigue, joint pain and others for several weeks after the disease has subsided. After 40 days of hospital discharge on follow-up, 4 patients complained of fatigue and 2 complained of generalized body ache on the SOC arm as compared to one patient who complained of on and off breathlessness on PNB001+SOC arm (this patient required long term oxygen therapy during the trial). No patients in the PNB001+SOC arm complained of fatigue or body ache. This may be due to early blunting of the inflammatory response and early oxygen weaning from oxygen therapy.

### Strengths and limitations of this study

This randomized clinical study assessed the comparative efficacy and safety PNB001+SOC versus SOC using comprehensive clinical endpoints in moderate COVID-19 patients. We consider that our findings are of high significance as the screened patients underwent randomization, and the baseline characteristics were similar between the treatment arms. All assessed parameters showed significant benefit or favored PNB001+SOC arm.

There were a few limitations to the study. The total number of patients enrolled in the study was low, however the number was based on regulatory recommendation during approval. Study was designed as an open label study however since open label was the requirement from the regulatory agency and since endpoints were mostly objective it is unlikely to have biased the endpoint assessment. Three patients (one in treatment and two in control) had to be discontinued due to worsening of clinical condition while another 2 withdrew after randomization into the study, however, this was essential to comply with the protocol.

## CONCLUSION

PNB001 with standard of care showed significant clinical improvement in moderate COVID-19 patients when compared to standard of care. PNB001 was well tolerated by moderate COVID-19 patients.

**What is already known on this topic**

- Release of large amount of pro-inflammatory cytokines results in an aggressive acute respiratory distress syndrome, causing tissue damage leading to multiple organ failure, and unfavourably affecting prognosis of severe COVID-19.
- The NLR ratio is a predictive biomarker for COVID-19 infection.
- PNB001 is first in class potent CCK-A agonists and CCK-B antagonist, the unique mechanism of its action being its anti-inflammatory and immune stimulation properties.

**What this study adds**

- PNB001 in combination with standard of care showed significant clinical improvement as assessed by mean change in 8-point WHO ordinal scale when compared to standard of care by Day 14. Primary endpoint of mortality rate and all the secondary endpoints showed quantitatively more benefit with PNB001
- PNB001 100 mg thrice daily along with SOC was well tolerated and showed similar safety profile as the SOC arm.
- Anti-inflammatory activity of potent steroids was further improved by PNB001. The NLR, which assess the immune stimulating properties and is the main mechanism to fight COVID19 infection was significantly reduced by PNB001.

## Data Availability

The data that support the findings of this study are available from the corresponding author, upon reasonable request.

## Acknowledgements

The authors would like to thank study participants for their contribution in the study.

## Contributors

Eric Lattmann, Pradnya Bhalerao, ShashiBhushan BL, Neeta Nargundkar, Pornthip Lattmann, Sadasivan Pillai K, Balaram PN, all the authors, contributed to critical discussion of trial design, and conduct, review of trial data, and revisions of the manuscript.

## Ethical approval

This study (Clinical Trial Registry of India: CTRI/2020/10/028423) was conducted by Biosphere Clinical Research Pvt. Ltd. Thane, Maharashtra following approval by the Drugs Controller General of India. The study protocol was approved by independent ethics committee of B.J. Government medical college and Sassoon General Hospital (ECR/280/Inst/Maha/2013/RR-19), and by independent ethics committee of Bangalore Medical College and Research Institute (ECR/302/Inst/KA/2013/RR-20). The study conduct was initiated after receiving the approval from both the independent ethics committee. The safety updates/reports including reports of suspected unexpected serious adverse reactions, were submitted to the independent ethics committee during the study. All the patients provided written informed consent prior to study enrolment.

## Data Sharing Statement

The lead author (Eric Lattmann) affirms that the manuscript is an honest, accurate, and transparent account of the study being reported; that no important aspects of the study have been omitted; and that any discrepancies from the study as originally planned (and, if relevant, registered) have been explained.

## REFERENCES

1. WHO. Coronavirus disease 2019 (COVID-19) Situation Report 52. Published online 2020. https://www.who.int/docs/default-source/coronaviruse/situation-reports/20200312-sitrep-52-covid-19.pdf?sfvrsn$=$e2bfc9c0_4

2. Ragab D, Salah Eldin H, Taeimah M, Khattab R, Salem R. The COVID-19 Cytokine Storm; What We Know So Far. Front Immunol. 2020;11. doi:10.3389/fimmu.2020.01446

3. Zhang C, Wu Z, Li J-W, Zhao H, Wang G-Q. Cytokine release syndrome in severe COVID-19: interleukin-6 receptor antagonist tocilizumab may be the key to reduce mortality. Int J Antimicrob Agents. 2020;55(5):105954. doi:10.1016/j.ijantimicag.2020.105954

4. Ruan Q, Yang K, Wang W, Jiang L, Song J. Clinical predictors of mortality due to COVID-19 based on an analysis of data of 150 patients from Wuhan, China. Intensive Care Med. 2020;46(5):846–848. doi:10.1007/s00134-020-05991-x

5. Tang Y, Liu J, Zhang D, Xu Z, Ji J, Wen C. Cytokine Storm in COVID-19: The Current Evidence and Treatment Strategies. Front Immunol. 2020;11:1708. Published 2020 Jul 10. doi:10.3389/fimmu.2020.01708.

6. Leisman DE, Ronner L, Pinotti R, et al. Cytokine elevation in severe and critical COVID- 19: a rapid systematic review, meta-analysis, and comparison with other inflammatory syndromes. Lancet Respir Med. 2020;8(12):1233–1244. doi:10.1016/S2213-2600(20)30404-5

7. Xia X, Wu J, Liu H, Xia H, Jia B, Huang W. Epidemiological and initial clinical characteristics of patients with family aggregation of COVID-19. J Clin Virol. 2020;127:104360. doi:10.1016/j.jcv.2020.104360

8. Pimentel GD, Dela Vega MCM, Laviano A. High neutrophil to lymphocyte ratio as a prognostic marker in COVID-19 patients. Clin Nutr Espen. 2020;40:101–102. doi:10.1016/j.clnesp.2020.08.004

9. LagunasCRangel FA. NeutrophilCtoClymphocyte ratio and lymphocyteCtoCCCreactive protein ratio in patients with severe coronavirus disease 2019 (COVIDC19): A metaCanalysis. J Med Virol. Published online April 8, 2020. doi:10.1002/jmv.25819

10. Fei F, Smith JA, Cao L. Clinical laboratory characteristics in patients with suspected COVID-19: One single-institution experience. J Med Virol. 2021;93(3):1665–1671. doi: https://doi.org/10.1002/jmv.26527

11. Veras FP, Pontelli MC, Silva CM, et al. SARS-CoV-2–triggered neutrophil extracellular traps mediate COVID-19 pathology. J Exp Med. 2020;217(e20201129). doi:10.1084/jem.20201129

12. Lattmann E, Sattayasai J, Narayanan R, et al. Cholecystokinin-2/gastrin antagonists: 5- hydroxy-5-aryl-pyrrol-2-ones as anti-inflammatory analgesics for the treatment of inflammatory bowel disease †The authors declare no competing interests. ‡Electronic supplementary information (ESI) available. See DOI: 10.1039/c6md00707d. MedChemComm. 2017;8(3):680–685. doi:10.1039/c6md00707d

13. Lattmann E, Sattayasai J. Cannabis, Cannabinoids and Tinnitus. J Pharmacol Drug Metab. 2014;1(1):1.

14. PNB Vesper gets DCGI nod to test proprietary drug PNB 001 in COVID-19 patients. Express Pharma. Published September 11, 2020. Accessed February 28, 2021. https://www.expresspharma.in/covid19-updates/pnb-vesper-gets-dcgi-nod-to-test-proprietary-drug-pnb-001-in-covid-19-patients/

15. Lattmann E, Lattmann P. A placebo-controlled, randomized, double blind, single ascending dose study to assess safety, tolerability, pharmacokinetics and pharmacodynamics of PNB-001 in healthy adult male subjects. Int J Dent Med Sci Res. 2021;3(1):130–144. doi: 10.35629/5252-0301130144.

16. Balaram PN, Lattmann P. A placebo-controlled, randomized, double blind, multiple ascending dose (MAD), multiple cohort, phase I study to assess safety, tolerability, pharmacokinetics of PNB-001 (Baladol) Int J Dent Med Sci Res. 2021;3(1):263–279. doi: 10.35629/5252-0301263279.

17. Ader F. Protocol for the DisCoVeRy trial: multicentre, adaptive, randomised trial of the safety and efficacy of treatments for COVID-19 in hospitalised adults. BMJ Open. 2020;10(9). doi:10.1136/bmjopen-2020-041437

18. WHO R&D Blueprint: novel Coronavirus COVID-19 Therapeutic Trial Synopsis. https://www.who.int/blueprint/priority-diseases/key-action/COVID-19_Treatment_Trial_Design_Master_Protocol_synopsis_Final_18022020.pdf

19. Natanegara F, Zariffa N, Buenconsejo J, et al. Statistical Opportunities to Accelerate Development for COVID-19 Therapeutics. Stat Biopharm Res. 2020;0(0):1–17. doi:10.1080/19466315.2020.1865195

20. Fernández-Ruiz M, López-Medrano F, Pérez-Jacoiste Asín MA, et al. Tocilizumab for the treatment of adult patients with severe COVID-19 pneumonia: A single-center cohort study. J Med Virol. 2021;93(2):831–842. doi:10.1002/jmv.26308

21. Beigel JH, Tomashek KM, Dodd LE, et al. Remdesivir for the Treatment of Covid-19 — Final Report. N Engl J Med. Published online May 22, 2020. doi:10.1056/NEJMoa2007764

22. Yang A-P, Liu J-P, Tao W-Q, Li H-M. The diagnostic and predictive role of NLR, d- NLR and PLR in COVID-19 patients. Int Immunopharmacol. 2020;84:106504. doi:10.1016/j.intimp.2020.106504

23. Upadhyay J, Tiwari N, Ansari MN. Role of inflammatory markers in corona virus disease (COVID-19) patients: A review. Exp Biol Med. 2020;245(15):1368–1375. doi:10.1177/1535370220939477

24. Vafadar Moradi E, Teimouri A, Rezaee R, et al. Increased age, neutrophil-to-lymphocyte ratio (NLR) and white blood cells count are associated with higher COVID-19 mortality. Am J Emerg Med. 2021;40:11–14. doi:10.1016/j.ajem.2020.12.003

25. Bosmans G, Shimizu Bassi G, Florens M, Gonzalez-Dominguez E, Matteoli G, Boeckxstaens GE. Cholinergic Modulation of Type 2 Immune Responses. Front Immunol. 2017;8:1873. doi:10.3389/fimmu.2017.01873

26. Yoo BB, Mazmanian SK. The Enteric Network: Interactions between the Immune and Nervous Systems of the Gut. Immunity. 2017;46(6):910–926. doi:10.1016/j.immuni.2017.05.011

27. Reardon C, Duncan GS, Brüstle A, et al. Lymphocyte-derived ACh regulates local innate but not adaptive immunity. Proc Natl Acad Sci. Published online January 7, 2013. doi:10.1073/pnas.1221655110

28. Eisner F, Martin EM, Küper MA, Raybould HE, Glatzle J. CCK1-Receptor Stimulation Protects Against Gut Mediator-Induced Lung Damage During Endotoxemia. Cell Physiol Biochem. 2013;32(6):1878–1890. doi:10.1159/000356644

29. Zhang C, Wu Z, Li J-W, Zhao H, Wang G-Q. Cytokine release syndrome in severe COVID-19: interleukin-6 receptor antagonist tocilizumab may be the key to reduce mortality. Int J Antimicrob Agents. 2020;55(5):105954. doi:10.1016/j.ijantimicag.2020.105954

30. Khadke S, Ahmed N, Ahmed N, et al. Harnessing the immune system to overcome cytokine storm and reduce viral load in COVID-19: a review of the phases of illness and therapeutic agents. Virol J. 2020;17(1):154. doi:10.1186/s12985-020-01415-w

31. Lattmann E, Russell ST, Singh M, et al. N□substituted 5□hydroxy□pyrrol□2□ones based cholecystokinin□2 antagonists as experimental anticancer agents for the treatment of lung cancer. MOJ Drug Des Develop Ther. 2018;2(4):180□189. DOI: 10.15406/mojddt.2018.02.00045.

32. Pimentel GD, Dela Vega MCM, Laviano A. High neutrophil to lymphocyte ratio as a prognostic marker in COVID-19 patients. Clin Nutr ESPEN. 2020;40:101–102. doi:10.1016/j.clnesp.2020.08.004

