## Supplementary Table 1 for "Randomized, Comparative, Clinical Trial to Evaluate Efficacy and Safety of PNB001 in Moderate COVID-19 Patients"

**Supplementary Table 1: Summary of Demographics and Baseline Characteristics**

| **Characteristics** | **Statistics** | **PNB001 (N=20)** | **Standard of Care (N=20)** | **All (N=40)** | **p-value** |
| --- | --- | --- | --- | --- | --- |
| Age | n | 20 | 20 | 40 | 0.8426 |
|  | Mean (SD) | 52.10 (12.83) | 52.80 (8.98) | 52.45 (10.94) |  |
|  | Median  (Min – Max) | 54.0  (30.0, 73.0) | 56.50  (34.0, 64.0) | 55.0  (30.0, 73.0) |  |
| Height | n | 20 | 20 | 40 | 0.4762 |
|  | Mean (SD) | 158.30 (6.97) | 156.0 (7.94) | 157.45 (7.42) |  |
|  | Median  (Min – Max) | 156.0  (150.0, 176.0) | 156.50  (146.0, 179.0) | 156.50  (146.0, 179.0) |  |
| Weight | n | 20 | 20 | 40 | 0.1592 |
|  | Mean (SD) | 61.90 (13.94) | 56.75 (7.94) | 59.33 (11.49) |  |
|  | Median  (Min – Max) | 57.0  (45.0, 95.0) | 58.50  (45.0, 70.0) | 58.0  (45.0, 95.0) |  |
| Sex | Male | 14 (70.00%) | 12 (60.00%) | 26 (65.00%) | 0.5073 |
|  | Female | 06 (30.00%) | 08 (40.00%) | 14 (35.00%) |  |
| Race | Indian | 20 (100%) | 20 (100%) | 40 (100%) | - |
| **Medical History** | | | | | |
| Diabetes Mellitus | | 04(20.00%) | 03 (15.00%) | 07 (17.50%) | - |
| Asthma | | 00 (00.00%) | 01 (05.00%) | 01 (02.50%) | - |
| Hypertension | | 02 (10.00%) | 04 (20.00%) | 06 (15.00%) | - |
| Hypothyroidism | | 01 (05.00%) | 00 (00.00%) | 01 (02.50%) | - |
| DVT | | 02 (10.00%) | 00 (00.00%) | 02 (05.00%) | - |
| Pulmonary embolism | | 01 (05.00%) | 00 (00.00%) | 01 (02.50%) | - |
| Obesity | | 00 (00.00%) | 01 (05.00%) | 01 (02.50%) | - |
| Hyperglycaemia | | 01 (05.00%) | 00 (00.00%) | 01 (02.50%) | - |
| SOC = Standard of Care | | | | | |
