## Supplementary Table 2 for "Randomized, Comparative, Clinical Trial to Evaluate Efficacy and Safety of PNB001 in Moderate COVID-19 Patients"

**Supplementary Table 2: Ordinal Scale for Clinical Improvement**

| **Patient State** | **Descriptor** | **Score** |
| --- | --- | --- |
| Uninfected | No clinical or virological evidence of infection | 0 |
| Ambulatory | No limitation of activities | 1 |
|  | Limitation of activities | 2 |
| Hospitalized  Mild disease | No oxygen therapy | 3 |
|  | Oxygen therapy by mask or nasal prongs | 4 |
| Hospitalized  Severe disease | Non-invasive ventilation or high-flow oxygen | 5 |
|  | Intubation and medical ventilation | 6 |
|  | Ventilation + additional organ support – pressors, RRT, ECMO | 7 |
| Dead | Death | 8 |
| ECMO=extracorporeal membrane oxygenation; RRT=renal replacement therapy | | |
