## Supplementary Table 3 for "Randomized, Comparative, Clinical Trial to Evaluate Efficacy and Safety of PNB001 in Moderate COVID-19 Patients"

**Supplementary Table 3: Summary Showing Different Methods of Oxygen Administration**

| **Visits** | **Scale** | **PNB001 (N=20)**  **n (%)** | **Standard of Care**  **(N=20)**  **n (%)** |
| --- | --- | --- | --- |
| **Baseline**  **(Day -1)** | 1-Off O2 | 00 (00.00%) | 00 (00.00%) |
|  | 2-O2 Mask/Nasal Prongs/Face Mask | 12 (60.00%) | 10 (50.00%) |
|  | 3-NRBM | 05 (25.00%) | 08 (40.00%) |
|  | 4-NIV/HFNO2 | 03 (15.00%) | 02 (10.00%) |
| **Day 1** | 1-Off O2 | 00 (00.00%) | 00 (00.00%) |
|  | 2-O2 Mask/Nasal Prongs/Face Mask | 12 (60.00%) | 10 (50.00%) |
|  | 3-NRBM | 06 (30.00%) | 06 (30.00%) |
|  | 4-NIV | 02 (10.00%) | 04 (20.00%) |
| **Day 2** | 1-Off O2 | 00 (00.00%) | 02 (10.00%) |
|  | 2-O2 Mask/Nasal Prongs/Face Mask | 11 (55.00%) | 11 (55.00%) |
|  | 3-NRBM | 07 (35.00%) | 03 (15.00%) |
|  | 4-NIV | 02 (10.00%) | 04 (20.00%) |
| **Day 3** | 1-Off O2 | 02 (10.00%) | 03 (15.00%) |
|  | 2-O2 Mask/Nasal Prongs/Face Mask | 10 (50.00%) | 13 (65.00%) |
|  | 3-NRBM | 06 (30.00%) | 00 (00.00%) |
|  | 4-NIV | 02 (10.00%) | 04 (20.00%) |
| **Day 4** | 1-Off O2 | 05 (26.32%) | 04 (20.00%) |
|  | 2-O2 Mask/Nasal Prongs/Face Mask | 09 (47.37%) | 11 (55.00%) |
|  | 3-NRBM | 03 (15.79%) | 00 (00.00%) |
|  | 4-NIV | 02 (10.53%) | 05 (25.00%) |
| **Day 5** | 1-Off O2 | 08 (42.11%) | 05 (26.32%) |
|  | 2-O2 Mask/Nasal Prongs/Face Mask | 07 (36.84%) | 09 (47.37%) |
|  | 3-NRBM | 02 (10.53%) | 00 (00.00%) |
|  | 4-NIV | 02 (10.53%) | 05 (26.32%) |
| **Day 6** | 1-Off O2 | 10 (52.63%) | 05 (27.78%) |
|  | 2-O2 Mask/Nasal Prongs/Face Mask | 06 (31.58%) | 08 (44.44%) |
|  | 3-NRBM | 01 (05.26%) | 01 (05.56%) |
|  | 4-NIV | 02 (10.53%) | 04 (22.22%) |
| **Day 7** | 1-Off O2 | 14 (73.68%) | 07 (38.89%) |
|  | 2-O2 Mask/Nasal Prongs/Face Mask | 02 (10.53%) | 06 (33.33%) |
|  | 3-NRBM | 01 (05.26%) | 02 (11.11%) |
|  | 4-NIV | 02 (10.53%) | 03 (16.67%) |
| **Day 8** | 1-Off O2 | 15 (83.33%) | 10 (55.56%) |
|  | 2-O2 Mask/Nasal Prongs/Face Mask | 02 (11.11%) | 03 (16.67%) |
|  | 3-NRBM | 00 (00.00%) | 03 (16.67%) |
|  | 4-NIV | 01 (05.56%) | 02 (11.11%) |
| **Day 9** | 1-Off O2 | 15 (83.33%) | 12 (70.59%) |
|  | 2-O2 Mask/Nasal Prongs/Face Mask | 02 (11.11%) | 01 (05.88%) |
|  | 3-NRBM | 00 (00.00%) | 04 (23.53%) |
|  | 4-NIV | 01 (05.56%) | 00 (0.00%) |
| **Day 10** | 1-Off O2 | 16 (88.89%) | 12 (70.59%) |
|  | 2-O2 Mask/Nasal Prongs/Face Mask | 01 (05.56%) | 01 (05.88%) |
|  | 3-NRBM | 00 (00.00%) | 04 (23.53%) |
|  | 4-NIV | 01 (05.56%) | 00 (00.00%) |
| **Day 11** | 1-Off O2 | 17 (94.44%) | 12 (70.59%) |
|  | 2-O2 Mask/Nasal Prongs/Face Mask | 00 (00.00%) | 02 (11.76%) |
|  | 3-NRBM | 00 (00.00%) | 03 (17.65%) |
|  | 4-NIV | 01 (05.56%) | 00 (00.00%) |
| **Day 12** | 1-Off O2 | 16 (88.89%) | 12 (70.59%) |
|  | 2-O2 Mask/Nasal Prongs/Face Mask | 01 (05.56%) | 03 (17.65%) |
|  | 3-NRBM | 01 (05.56%) | 02 (11.76%) |
|  | 4-NIV | 00 (00.00%) | 00 (00.00%) |
| **Day 13** | 1-Off O2 | 16 (88.89%) | 13 (76.47%) |
|  | 2-O2 Mask/Nasal Prongs/Face Mask | 01 (05.56%) | 02 (11.76%) |
|  | 3-NRBM | 01 (05.56%) | 02 (11.76%) |
|  | 4-NIV | 00 (00.00%) | 00 (00.00%) |
| **Day 14** | 1-Off O2 | 17 (94.44%) | 13 (76.47%) |
|  | 2-O2 Mask/Nasal Prongs/Face Mask | 01 (05.56%) | 02 (11.76%) |
|  | 3-NRBM | 00 (00.00%) | 02 (11.76%) |
|  | 4-NIV | 00 (00.00%) | 00 (00.00%) |
