## Supplementary Table 4 for "Randomized, Comparative, Clinical Trial to Evaluate Efficacy and Safety of PNB001 in Moderate COVID-19 Patients"

**Supplementary Table 4: Summary of Exploratory Efficacy Endpoints**

| **Days** | **Statistics** | **PNB001** | **Standard of Care** | **P-value** |
| --- | --- | --- | --- | --- |
| **IL-6 (pg/ml))** | | | | |
| **Baseline**  **(Day -1)** | n | 17 | 16 | - |
|  | Mean (SD) | 67.82 (87.93) | 61.96 (63.56) | - |
|  | 95% CI of Mean | 22.61-113.0 | 28.09-95.83 | - |
|  | Median (Min, Max) | 30.27 (4.000, 312.3) | 33.22 (5.760, 188.3) | - |
| **End of Study**  **(Day 14)** | N | 15 | 13 | - |
|  | Mean (SD) | 23.25 (30.75) | 22.41 (19.31) | - |
|  | 95% CI of Mean | 6.22-40.28 | 10.74-34.08 | - |
|  | Median (Min, Max) | 13.08 (2.270, 112.8) | 19.45 (2.510, 62.90) | - |
| **Change from baseline to End of Study** | n | 15 | 13 | - |
|  | Mean (SD) | 49.86 (96.74) | 38.64 (51.87) | - |
|  | 95% CI of Mean | -3.717, 103.4 | 7.292, 69.98 | - |
|  | Median (Min, Max) | 25.98 (-105.7, 268.9) | 24.05 (-17.12, 125.4) | - |
|  | P-value within Treatment | 0.0151 | 0.0067 | - |
|  | P-value between Treatment | - | - | 0.7471 |
| **Neutrophil/Lymphocyte Ratio** | | | | |
| **Baseline**  **(Day -1)** | n | 15 | 18 | - |
|  | Mean (SD) | 10.36 (4.846) | 12.27 (10.21) | - |
|  | 95% CI of Mean | 7.675-13.04 | 7.197-17.35 | - |
|  | Median (Min, Max) | 8.800 (6.052, 24.000) | 10.13 (4.460, 48.000) | - |
| **End of Study**  **(Day 14)** | N | 13 | 15 | - |
|  | Mean (SD) | 3.127 (1.586) | 6.438 (4.401) | - |
|  | 95% CI of Mean | 2.168-4.085 | 4.000-8.875 | - |
|  | Median (Min, Max) | 2.913 (1.019, 6.814) | 6.062 (2.186, 18.40) | - |
| **Change from baseline to End of Study** | n | 13 | 15 | - |
|  | Mean (SD) | 6.435 (4.846) | 6.032 (12.13) | - |
|  | 95% CI of Mean | 3.507, 9.36644 | -0.6872, 12.75 | - |
|  | Median (Min, Max) | 4.880 (1.986, 20.91) | 2.243 (-6.113, 45.04) | - |
|  | P-value within Treatment | 0.0001 | 0.0148 | - |
|  | P-value between Treatment | - | - | 0.1971 |
| **Erythrocytes Sedimentation Rate (mm at 1 h)** | | | | |
| **Baseline**  **(Day -1)** | n | 20 | 19 | - |
|  | Mean (SD) | 44.05 (21.83) | 39.84 (25.44) | - |
|  | 95% CI of Mean | 33.83-54.27 | 27.58-52.10 | - |
|  | Median (Min, Max) | 36.50 (16.00, 90.00) | 31.00 (10.00, 94.00) | - |
| **End of Study**  **(Day 14)** | N | 18 | 16 | - |
|  | Mean (SD) | 27.17 (18.73) | 21.63 (14.76) | - |
|  | 95% CI of Mean | 17.85-36.48 | 13.76-29.49 | - |
|  | Median (Min, Max) | 20.50 (8.00, 90.00) | 19.00 (11.00, 75.00) | - |
| **Change from baseline to End of Study** | n | 18 | 16 | - |
|  | Mean (SD) | 17.89 (29.48) | 14.38 (28.12) | - |
|  | 95% CI of Mean | 3.23, 32.55 | -0.6070, 29.36 | - |
|  | Median (Min, Max) | 19.50 (-60.00, 64.00) | 8.00 (-41.00, 70.00) | - |
|  | P-value within Treatment | 0.0065 | 0.0143 | - |
|  | P-value between Treatment | - | - | 0.3004 |
| **C-Reactive Protein (mg/l)** | | | | |
| **Baseline**  **(Day -1)** | n | 20 | 19 | - |
|  | Mean (SD) | 44.05 (21.83) | 39.84 (25.44) | - |
|  | 95% CI of Mean | 33.83-54.27 | 27.58-52.10 | - |
|  | Median (Min, Max) | 36.50 (16.00, 90.00) | 31.00 (10.00, 94.00) | - |
| **End of Study**  **(Day 14)** | N | 18 | 16 | - |
|  | Mean (SD) | 27.17 (18.73) | 21.63 (14.76) | - |
|  | 95% CI of Mean | 17.85-36.48 | 13.76-29.49 | - |
|  | Median (Min, Max) | 20.50 (8.00, 90.00) | 19.00 (11.00, 75.00) | - |
| **Change from baseline to End of Study** | n | 18 | 16 | - |
|  | Mean (SD) | 17.89 (29.48) | 14.38 (28.12) | - |
|  | 95% CI of Mean | 3.23, 32.55 | -0.6070, 29.36 | - |
|  | Median (Min, Max) | 19.50 (-60.00, 64.00) | 8.00 (-41.00, 70.00) | - |
|  | P-value within Treatment | 0.0065 | 0.0143 | - |
|  | P-value between Treatment | - | - | 0.3004 |
| **Platelet/Lymphocyte Ratio** | | | | |
| **Baseline**  **(Day -1)** | n | 14 | 12 | - |
|  | Mean (SD) | 328.6 (178.0) | 345.3 (232.2) | - |
|  | 95% CI of Mean | 225.8-431.3 | 216.7-473.9 | - |
|  | Median (Min, Max) | 306.3 (35.94, 651.7) | 315.9 (78.35, 876.8) | - |
| **End of Study**  **(Day 14)** | N | 12 | 12 | - |
|  | Mean (SD) | 114.6 (52.76) | 181.4 (107.2) | - |
|  | 95% CI of Mean | 81.07-148.1 | 113.3-249.5 | - |
|  | Median (Min, Max) | 108.6 (35.21, 214.6) | 134.0 (95.63, 447.9) | - |
| **Change from baseline to End of Study** | n | 12 | 12 | - |
|  | Mean (SD) | 206.4 (180.4) | 135.0 (231.0) | - |
|  | 95% CI of Mean | 229.4 (-70.44, 533.6) | 77.30 (-103.6, 779.7) | - |
|  | Median (Min, Max) | 0.0011 | 0.0340 | - |
|  | P-value within Treatment | - | - | 0.4078 |
|  | P-value between Treatment | 14 | 12 | - |
